# Aetiologies of chronic cough diagnosed using a pathophysiologic diagnostic procedure and their treatment outcomes

**DOI:** 10.1101/2021.02.19.21252115

**Authors:** Johsuke Hara, Masaki Fujimura, Masahide Yasui, Reiko Takeda, Noriyuki Ohkura

**Affiliations:** Department of Respiratory Medicine, Kanazawa University Hospital, Kanazawa, Ishikawa, Japan, 13-1, Takara-machi, Kanazawa, Ishikawa, 920-8641, Mail; Department of Respiratory Medicine, National Hospital Organization Nanao Hospital 8 bu 3-1 Matsuto-machi, Nanao, Ishikawa 926-8531; Laboratory Department, National Hospital Organization Nanao Hospital 8 bu 3-1 Matsuto-machi, Nanao, Ishikawa 926-8531

## Abstract

**Background:** We previously elucidated the fundamental pathophysiological features of various aetiologies of chronic cough, such as cough variant asthma, atopic cough, and sinobronchial syndrome. We also established a pathophysiological diagnostic procedure for aetiology identification. In this study, we aimed to disclose the aetiologies of chronic cough using the pathophysiological diagnostic procedure and to determine the outcomes of treatment administered on the basis of aetiology.

**Patients and Methods:** We retrospectively reviewed the medical records of patients with chronic cough who visited our cough specialty clinic from September 2013 to August 2018 and analyzed the pathophysiological diagnostic procedure-based aetiologies and corresponding treatments. The pathophysiological diagnostic procedure included the capsaicin cough test, methacholine cough test, bronchial reversibility test, bronchial responsiveness test, chest and sinus CT, and sputum examinations.

**Results:** Initially, 303 patients were selected, and 300 patients underwent the diagnostic procedure. Aetiologies of chronic cough were diagnosed in 297 patients (99.0%). In the other three patients (1.0%), all results of the diagnostic procedure were within normal limits; their aetiologies were evaluated using the therapeutic diagnostic procedure. Tweleve patients discontinued follow-up before completing treatment. Of the 291 remaining patients, cough resolved completely in 283 patients. The median time required for the complete resolution of cough was 5.0 weeks (95% CI 4.3∼5.7 weeks).

**Conclusions:** The pathophysiological diagnostic procedure can lead to rapid and objective diagnosis of causes of chronic cough, which leads to superior treatment results compared to therapeutic diagnostic procedure.

**Trial registration number:** UMIN ID: UMIN000018679

**Key Messages:** Our pathophysiological diagnostic procedure can lead to rapid and objective diagnosis of the aetiology of chronic cough, leading to superior treatment results compared to those of therapeutic diagnostic procedures.

## Introduction

Major aetiologies of chronic cough (CC) in Western countries are gastroesophageal reflux (GER), upper airway cough syndrome (UACS, rhinitis/sinusitis), asthmatic cough (cough variant asthma: CVA), and non-asthmatic eosinophilic bronchitis (NAEB) ^1,2^. However, CVA, atopic cough (AC), and sinobronchial syndrome (SBS) are the major causes of CC in Japan ^3^. Additionally, eosinophilic airway disorders presenting with chronic non-productive cough in Western countries include CVA and NAEB, while those in Japan include CVA and AC. The differences in the concept of eosinophilic airway disorder may have originated from differences in the criteria for the diagnosis of CVA between Japan and Western countries.

It is important to recognize that strong bronchodilators, such as β2 agonists, have no effects on peripheral cough receptor sensitivity ^4^ or the central cough center ^5^. In other words, bronchodilators basically have no antitussive effects. In 1979, Corrao et al. ^6^ reported six patients with CC relieved using oral bronchodilators (β2-agonists or theophylline); bronchial responsiveness to inhaled methacholine was slightly higher in these patients than in normal subjects. On this basis, we, cough specialists in Japan, have recognized CVA as a bronchodilator-responsive CC, while cough specialists in Western countries have recognized CVA as a CC associated with bronchial hyperresponsiveness.

Long-term series of studies conducted by our group and others have revealed the fundamental pathophysiologic features of CVA, AC, and SBS. Although some key features of CVA recognized in Japan and Western countries are similar (precursors of typical asthma, chronic airflow limitation in some patients), there is a key difference: In Japan, eosinophilic broncho-bronchiolitis with heightened cough response to bronchial smooth muscle contraction (smooth muscle cough hypersensitivity, SCH) occurs. In Western countries, eosinophilic broncho-bronchiolitis with bronchial hyperresponsiveness and cough-reflex hypersensitivity is present.

In the 1980s, we evaluated for bronchodilator-responsive non-productive cough, which is the key criterion for diagnosis of CVA according to Corrao et al. ^6^. In this process, we found many patients with chronic non-productive cough resistant to bronchodilator therapy, which led us to propose AC as a new disease entity to carry out our series of clinical and experimental studies on AC. Second-generation histamine H1 antagonists and inhaled and oral corticosteroids ^7-11^, but not leukotriene receptor antagonists ^12^, are effective against AC. The fundamental features of AC are as follows: 1) presence of eosinophilic tracheobronchitis with cough-reflex hypersensitivity (epithelial cough hypersensitivity, ECH); 2) absence of precursors of typical asthma, and 3) absence of chronic airflow limitation.

Currently, diagnostic procedures used to determine the aetiology of CC are mostly based on the efficacy of specific treatment against cough. There are concerns regarding the efficiency of these therapeutic diagnostic procedures, such as the following:

1. False positivity due to low specificity of specific therapies
2. False positivity due to spontaneous cough relief
3. False negativity due to insufficiency of treatment potency
4. False negativity due to treatment resistance (difficult-to-treat or severe condition)
5. False negativity because of the presence of two or more cough aetiologies
6. Differences in cough treatment response evaluation criteria between studies

β2-agonists—and probably leukotriene receptor antagonists ^12^—are specific treatments for CVA, and proton-pump inhibitors for GER-associated cough. However, histamine H1 antagonists are effective for the treatment of AC and UACS. Inhaled corticosteroids (ICSs) are effective against CVA, AC, and NAEB. In addition to the low specificity of treatment, spontaneous relief in cough, insufficient treatment, and difficulty in treatment lead to the misdiagnosis or lack of diagnosis of etiology (idiopathic?). A recent report indicated that drug doses were not designed to produce antitussive effects in many studies ^13^. Furthermore, differences in cough treatment response evaluation criteria between studies result in different in cough aetiologies among studies. However, accumulating evidence on the mechanisms of cough and the pathophysiology of CVA and AC may enable us to advance from therapeutic diagnosis to pathophysiologic diagnosis.

We have established a pathophysiological diagnostic procedure for determination of the aetiologies of CC. The procedure involves the capsaicin cough test, methacholine cough test, bronchial reversibility test, bronchial responsiveness test, chest and sinus CT, and sputum examinations. This study was designed to disclose aetiologies of CC, as determined using the pathophysiological diagnostic procedure, and to assess the outcomes of treatment administered on the basis of aetiology.

## Patients and methods

In this retrospective observational cohort study, patients who visited our cough specialty clinic for the diagnosis and treatment of CC from September 2013 to August 2018 were included. Data on the aetiology of CC, as determined using the pathophysiological diagnostic procedure and on treatment were obtained from medical records. All patients provided written informed consent to participate in the study. The study was conducted in accordance with the Declaration of Helsinki and was approved by the Ethical Review Board of National Hospital Organisation Nanao Hospital (UMIN ID: UMIN000018679).

The patients underwent the following examinations in the specified order within 4 days of the first visit, after providing informed consent: pulmonary function testing, cough-reflex sensitivity to inhaled capsaicin, bronchial reversibility, cough response to inhaled methacholine-induced bronchoconstriction, and bronchial responsiveness to inhaled methacholine. Forced vital capacity (FVC), forced expiratory volume in the first second (FEV1), and flow-volume curves were measured using a dry wedge spirometer (Chestac 11, Chest Co., Ltd., Tokyo, Japan). Spirometric tests and evaluation of data were conducted in accordance with the ATS/ERS task force guidelines ^14^. Bronchial reversibility was determined using spirometric tests performed before and 30 min after the inhalation of 50 µg procaterol.

### Methacholine inhalation protocol

Methacholine inhalation was performed using an Astograph (Jupiter 21; CHEST; Tokyo, Japan), according to the method described by Takishima et al. ^15^. Methacholine chloride (Wako Pure Chemical Industries, Ltd., Osaka, Japan) was diluted in phosphate-buffered saline solution (PBS) to obtain a series of concentrations with two-fold increase from 0.0195 mg/mL to 160 mg/mL. The PBS and methacholine solutions were inhaled for 1 min. Each subject, wearing a nose clip, was examined during quiet breathing in a sitting position.

### Assessment of cough response to bronchoconstriction induced by methacholine

Cough response to methacholine-induced bronchoconstriction was assessed as previously described ^16^. Spirometry was performed before methacholine inhalation and immediately after the respiratory resistance had increased two-fold, at which time, if FEV1 did not decrease to <90% of the baseline value, inhalation of methacholine was restarted at the same concentration. An observer counted coughs, and cough counts were recorded at intervals of <1 min (α min) before and for 30 min (total 30 + α min) following inhalation of methacholine, at which time the respiratory resistance and FEV1 were archived (Meth-C). When the Meth-C was ≥24 coughs/30 + α min, the cough response to methacholine-induced bronchoconstriction was judged to have increased (SCH) ^17^.

### Assessment of cough-reflex sensitivity to inhaled capsaicin

Cough-reflex sensitivity to capsaicin was measured using a method that we previously described ^18^. Capsaicin (Wako Pure Chemical Industries Ltd, Tokyo, Japan) (30.5 mg) was dissolved in Tween 80 (1 mL) and ethanol (1 mL) and then in normal saline (8 mL) to prepare a 3.05 mg/mL stock solution (1 × 10^−2^ mol/L), which was stored at −20 °C. This solution was diluted with physiological saline to obtain a series of concentrations with two-fold increase from 0.49 μmol/L to 1 mmol/L. Each subject initially inhaled the control solution (physiological saline), followed capsaicin solutions of progressively increasing concentrations. While wearing a nose clip, the subjects inhaled solutions through a Bennett twin nebulizer (3012–60 mL, Puritan-Bennett Company, Carlsbad, California, USA) for 15 s every 60 s. Increasing concentrations were administered until ≥5 coughs were elicited. The number of coughs was counted by a skilled medical technician for a total of 60 s, that is, 15 s of inhalation plus 45 s of observation with each concentration of capsaicin solution. The nebulizer output was 0.21 mL/min. The cough threshold was defined as the lowest concentration of capsaicin that elicited ≥5 coughs (C5). As there is a sex-based difference in the C5 ^19^ (female subjects: C5 ≤ 0.98 μmol/L; male subjects: C5 ≤ 3.9 μmol/L), cough sensitivity to inhaled capsaicin was judged to increase (ECH). The cut-off point was determined as follows in healthy subjects: geometric mean − 2 geometric standard deviations (unpublished data).

Our pathophysiologic diagnosis was based on the following criteria: CVA, increase in cough response to methacholine-induced airway smooth muscle contraction (SCH); AC, increase in cough-reflex sensitivity to inhaled capsaicin (ECH); a combination of CVA and AC, increase in cough response due to both airway smooth muscle contraction and cough-reflex sensitivity.

In the present study, when all the following criteria were met, the patient was diagnosed with CVA:

1. Isolated chronic non-productive cough lasting for ≥8 weeks
2. No history of wheezing or dyspnea and lack of adventitious lung sounds on physical examination
3. Increase in cough response to methacholine-induced bronchoconstriction

When all following criteria were met, the patient was diagnosed with AC:

1. Non-productive cough lasting for ≥8 weeks
2. Presence of one or more findings indicative of an atopic predisposition, including a history and/or complications of allergic diseases (excluding asthma), peripheral blood eosinophilia (> 6% or >400 cells/µL), increased total serum IgE level (>200 IU/mL), presence of IgE antibodies specific to aeroallergens, and positivity of allergen skin testing and/or presence of induced sputum eosinophilia (> 2.0%)
3. Increase in cough-reflex sensitivity to inhaled capsaicin

The diagnosis of SBS was in accordance to the following criteria:

1. Productive cough lasting for ≥8 weeks
2. One or more of the following:
  i. Symptoms such as postnasal drip and throat clearing
  ii. Signs such as mucous or mucopurulent secretion in the upper and middle pharynx and cobblestone appearance of the mucosa.
  iii. Fluid retention and/or mucosal thickening on sinus CT scan.
  iv. Increased neutrophil count in nasal secretions
3. Increased neutrophil count in spontaneous sputum.C
4. Cough relief upon treatment with 14-member macrolides

Treatment efficacy was evaluated at 2 months after initiation and was judged as effective when the productive cough diminished to half or less.

When all the following criteria were met, the patient was diagnosed with GER-associated cough:

1. Non-productive cough lasting for ≥8 weeks
2. GER based on the presence of one or more of the following characteristics:
  i. Digestive symptoms such as heartburn, epigastric or retrosternal discomfort, and sour taste
  ii. Oesophageal hernia or reflux esophagitis by esophagoscopy or reflux of barium up to the middle portion of the esophagus
  iii. Documentation of gastro-oesophageal reflux by 24-h ambulatory oesophageal pH monitoring
3. Relief of cough upon treatment with a combination of proton-pump inhibitors and acotiamide hydrochloride hydrate

Mucoid impaction of small bronchi (MISB) syndrome was diagnosed when the following criteria were met:

1. Productive cough lasting for ≥8 weeks
2. Increased eosinophil count in spontaneous sputum
3. Impaction of small bronchi and remarkable thickening of bronchial walls on chest CT

Although this clinical entity has not been established, we have encountered many patients who meet the above-mentioned criteria, and they were successfully treated with oral corticosteroids and antifungal drugs (in submission). The clinical features were as follows: 1) Cough was relieved upon short-term treatment with oral corticosteroids, but it relapsed soon after the termination of treatment (intractable); 2) Bacteria causing chronic airway infection were seldom detected in purulent sputum, and sputum eosinophil counts were increased; 3) long-term low-dose macrolide therapy was not effective; 4) MISB was mostly identified in the lower lobes; 5) fungi were seldom culturable in clinically available fungal growth media; and 6) a combination of oral corticosteroids and itraconazole was effective.

Although asthma may need to be excluded from chronic cough, patients with asthma complaining of isolated CC were included in this study because each chief physician could not detect asthma in a patient with no history of wheezing or dyspnea, no wheezes on auscultation, and no airflow limitation assessed by spirometry at the first presentation. The series of diagnostic procedures revealed that those patients had one or more characteristic features of asthma: bronchial hyperresponsiveness; bronchial reversibility, defined as a percentage increase of ≥ 12% and an absolute volume increase of 200 mL in FEV1; and/or wheezes on auscultation. Some physicians may diagnose asthma in those patients, and others will diagnose CVA. In the present study, we diagnosed cough-predominant asthma (CPA) in such subjects as a matter of convenience.

### Statistical analysis

Data are expressed as mean ± standard deviation (SD). The distribution of counts for the two groups (< 65 years and ≥ 65 years) was assessed using Pearson’s chi-square test. The Kruskal–Wallis H test was used to assess other inter-group variations. All comparisons were two-tailed, and p values < 0.05 were considered significant. Statistical analyses were performed using IBM SPSS Statistic23 (Japan IBM Co. Tokyo, Japan).

## Results

In total 303 patients (men, 122; women, 181; mean age, 57.0 ± 17.1 years) initially visited our clinic during the 5-year study period. Three patients did not undergo the complete pathophysiological diagnostic procedure. Among the remaining 300 patients, the causes of chronic cough were diagnosed in 297 patients (99.0%). In the other 3 patients (1.0%), all pathophysiological diagnostic procedure results were within normal limits, and therefore, the therapeutic diagnostic procedure was used; while 2 patients underwent the complete procedure, one patient dropped out (Figure 1). The frequency of each aetiology of CC, according to the findings of the pathophysiological diagnostic procedure, is shown in Table 1. Two or more causes were diagnosed in 218/300 patients (72.7%) (Table 1). CVA, AC, and SBS were the major causes of cough (Table 2). Higher number of causes were diagnosed in elderly patients (≥65 years of age) than in non-elderly patients (p =0.020) (Table 3).

**Figure 1.**
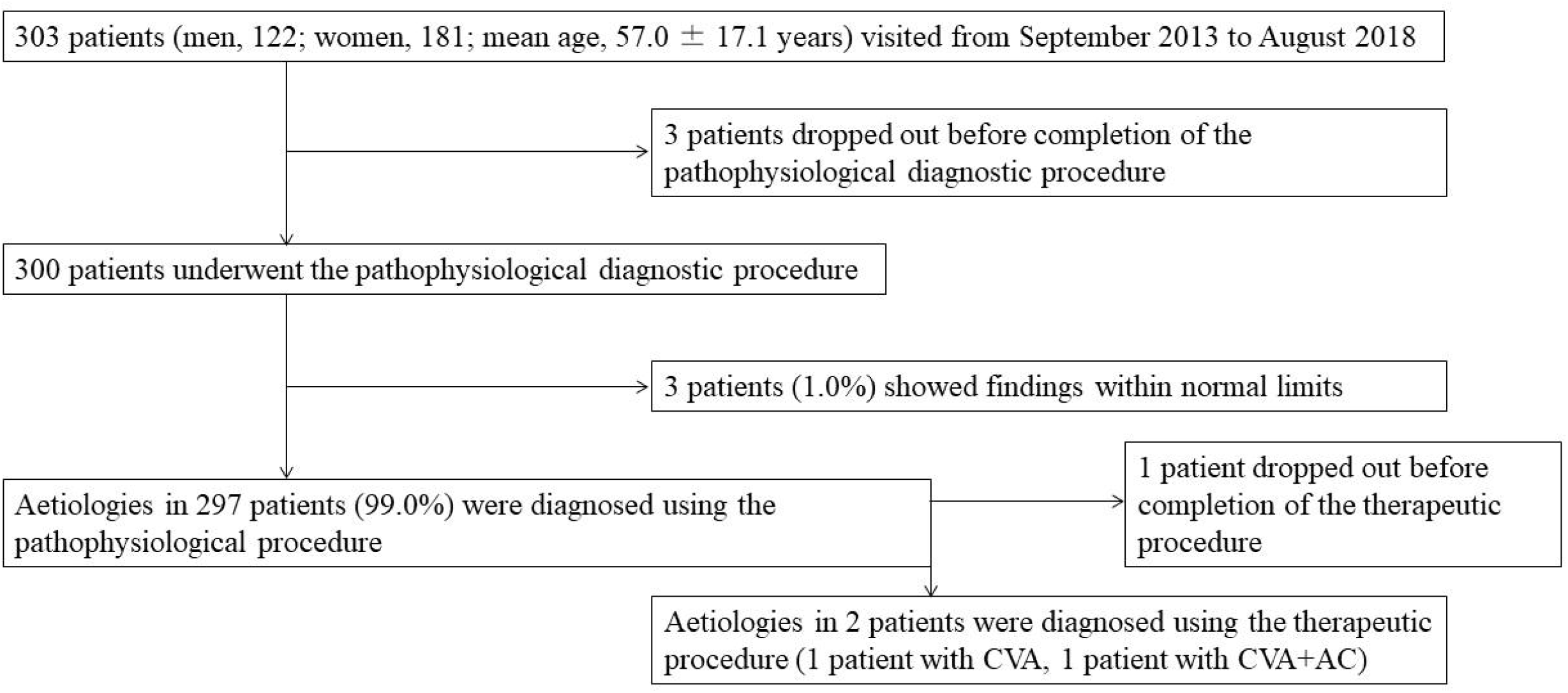
Details of patients who underwent the pathophysiological diagnostic procedure AC: atopic cough; CVA: cough variant asthma

**Table 1.**
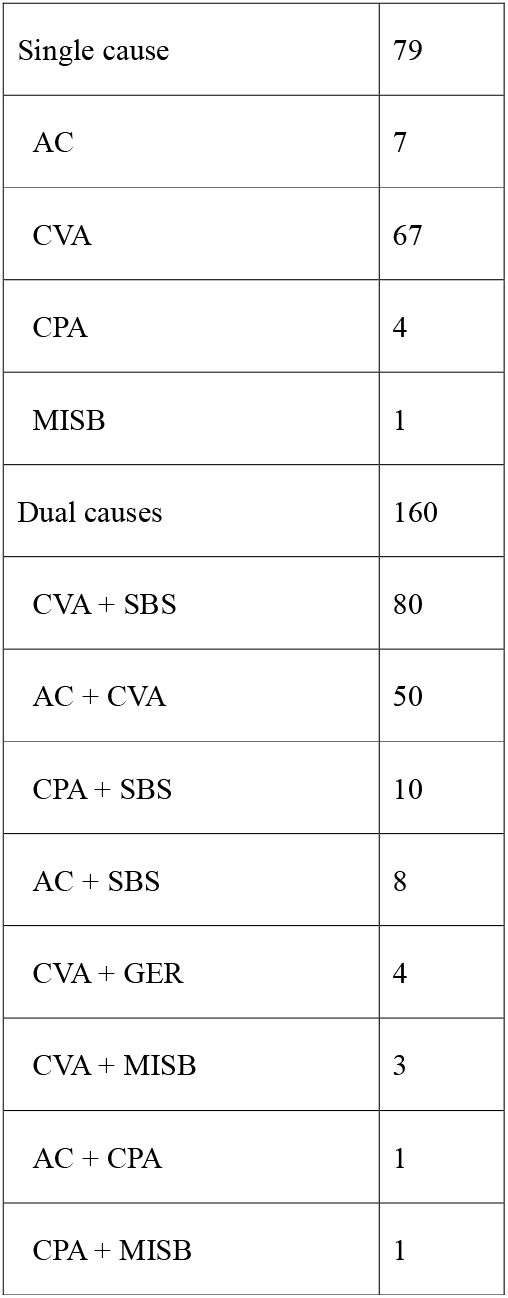
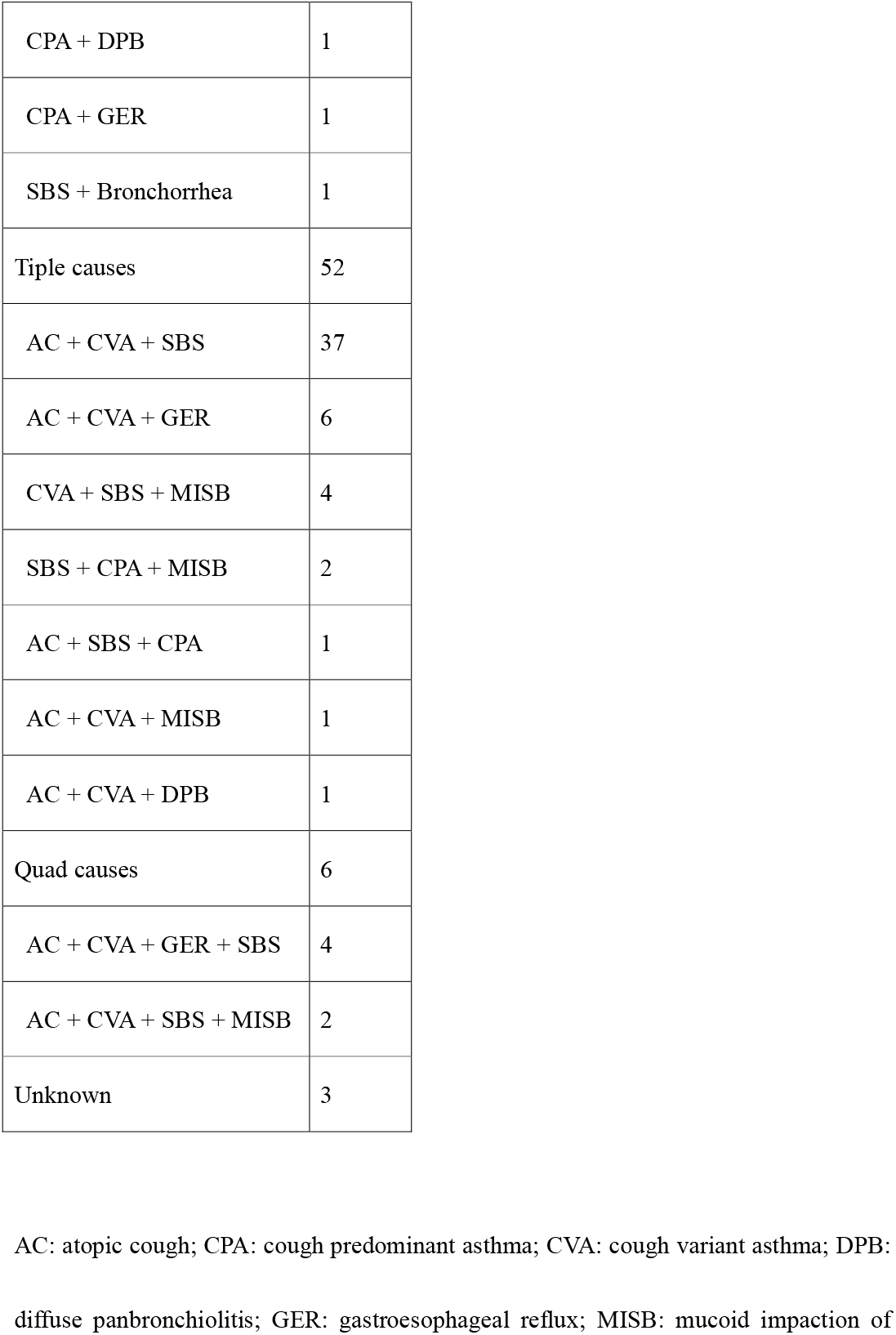
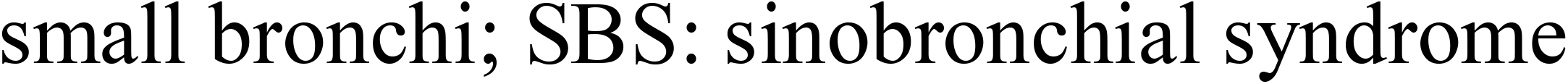
Aetiologies of chronic cough in 300 patients, as determined using the pathophysiological diagnostic procedure

|  |  |
| --- | --- |
| Single cause | 79 |
| AC | 7 |
| CVA | 67 |
| CPA | 4 |
| MISB | 1 |
| Dual causes | 160 |
| CVA + SBS | 80 |
| AC + CVA | 50 |
| CPA + SBS | 10 |
| AC + SBS | 8 |
| CVA + GER | 4 |
| CVA + MISB | 3 |
| AC + CPA | 1 |
| CPA + MISB | 1 |
| CPA + DPB | 1 |
| CPA + GER | 1 |
| SBS + Bronchorrhea | 1 |
| Tiple causes | 52 |
| AC + CVA + SBS | 37 |
| AC + CVA + GER | 6 |
| CVA + SBS + MISB | 4 |
| SBS + CPA + MISB | 2 |
| AC + SBS + CPA | 1 |
| AC + CVA + MISB | 1 |
| AC + CVA + DPB | 1 |
| Quad causes | 6 |
| AC + CVA + GER + SBS | 4 |
| AC + CVA + SBS + MISB | 2 |
| Unknown | 3 |

AC: atopic cough; CPA: cough predominant asthma; CVA: cough variant asthma; DPB: diffuse panbronchiolitis; GER: gastroesophageal reflux; MISB: mucoid impaction of
small bronchi; SBS: sinobronchial syndrome

**Table 2.** Numbers of patients diagnosed with major aetiologies of cough, as determined using the pathophysiological diagnostic procedure

|  |  |
| --- | --- |
| Major cough causes | N = 300 |
| CVA | 256 |
| SBS | 149 |
| AC | 116 |
| GER | 15 |
| MISB | 14 |
| DPB | 2 |

AC: atopic cough; CVA: cough variant asthma; DPB: diffuse panbronchiolitis; GER: gastroesophageal reflux; MISB: mucoid impaction of small bronchi; SBS: sinobronchial syndrome

**Table 3.**
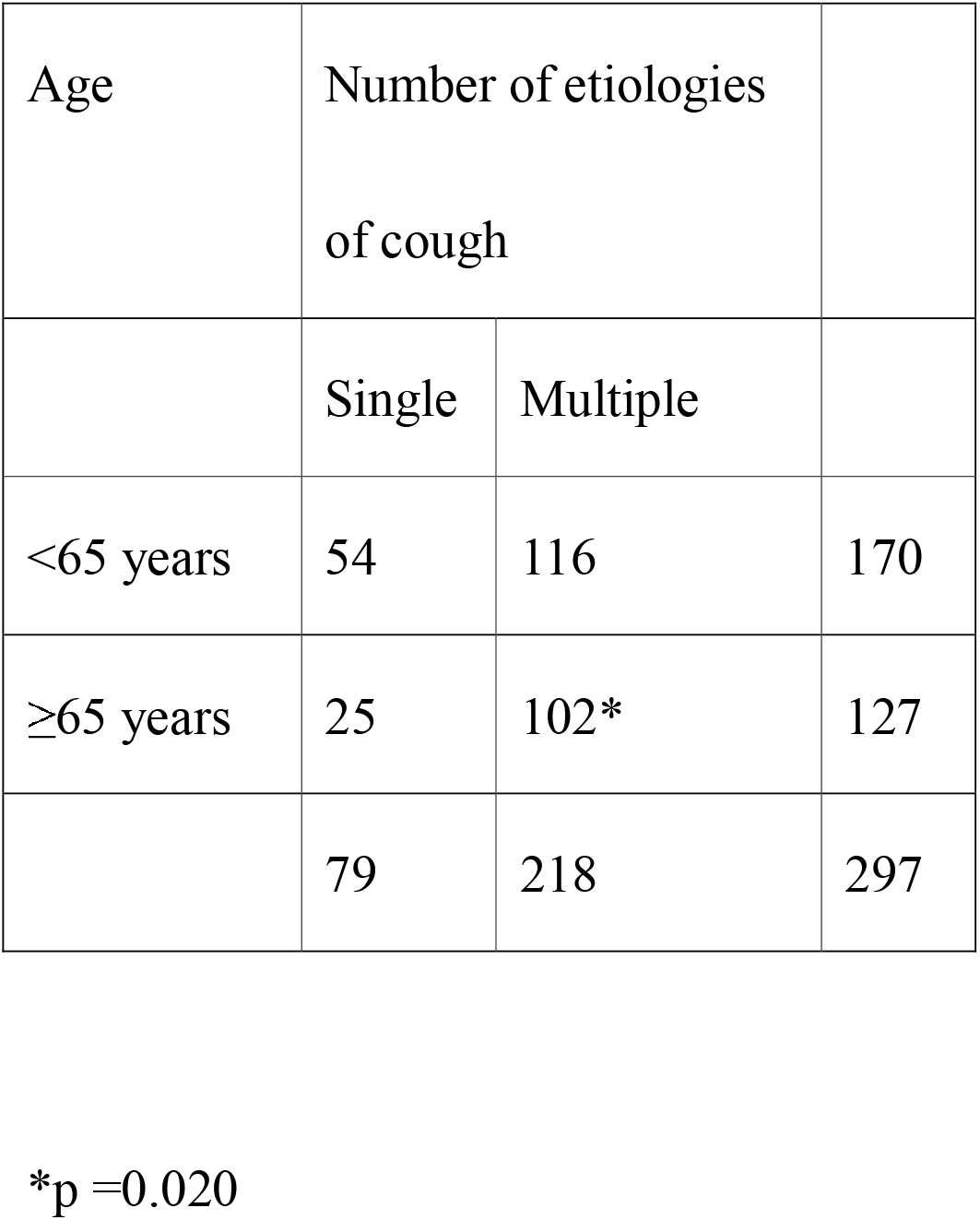
Age-related differences in the number of cough aetiologies

Twelve of the 303 patients dropped out before receiving sufficient medications. Of the remaining 291 patients, 283 (97.3%) experienced complete resolution of cough (Figure 2). In 8 patients (2.7%), the cough was not completely resolved. The intensities of remaining cough as indicated by visual analogue scale were as follows: 1/10 in 2 patients, 2/10 in 2 patients, 3/10 in 1 patient, 4/10 in 1 patient, and 5/10 in 2 patients. The median time required for the resolution of cough was 5.0 weeks (95% CI: 4.3–5.7 weeks) (Figure 3), and this time did not significantly differ depending on the number of aetiologies of cough (Figure 4). Frequencies of CC aetiologies diagnosed using the pathophysiological diagnostic procedure and the treatment response are shown in Table 4.

**Figure 2.**
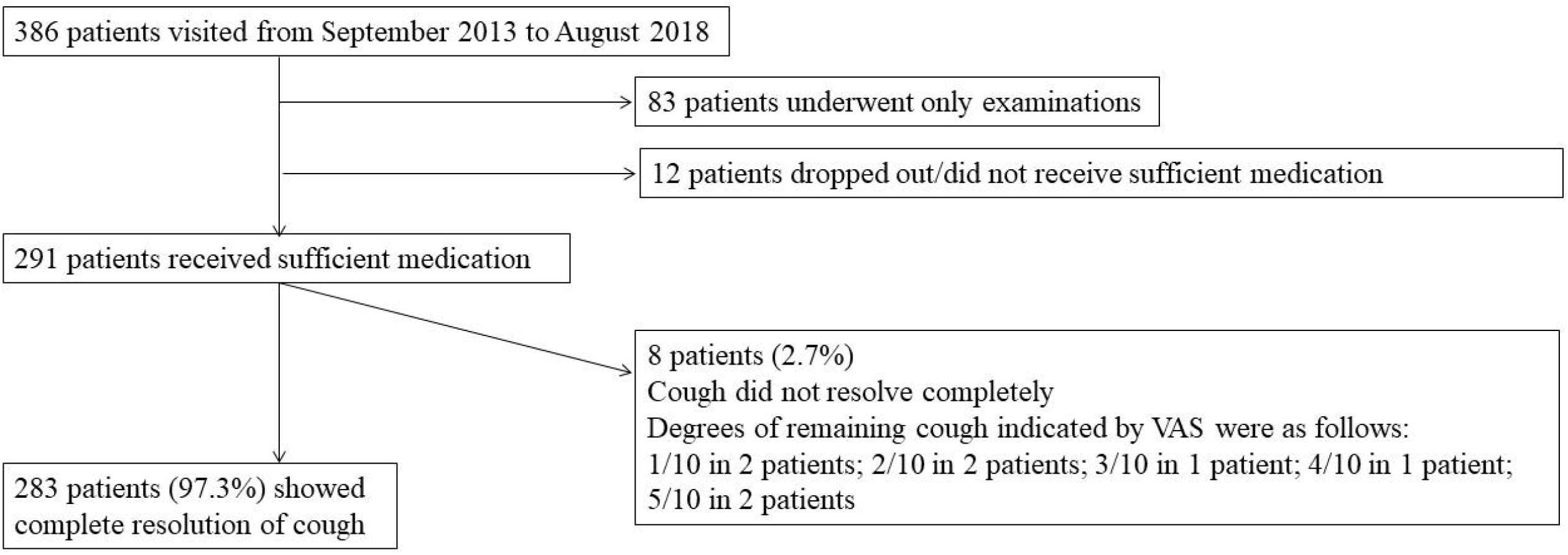
Treatment response for patients, in whom the aetiology of chronic cough was diagnosed using the pathophysiological diagnostic procedureVAS, visual analogue scale

**Figure 3.**
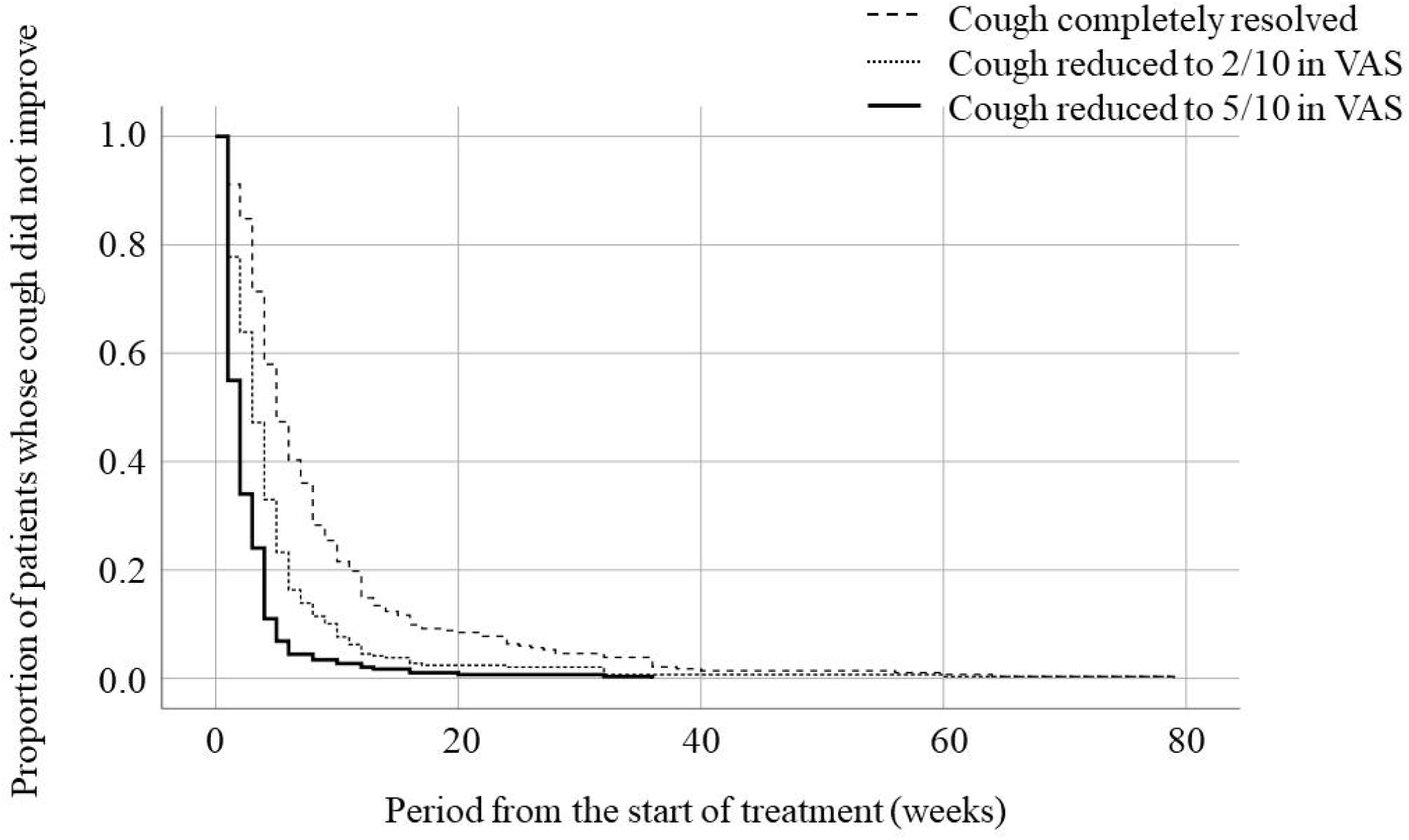
Time to complete cough resolution in 291 patients Dashed line: patients whose cough was completely resolved; dotted line: patients whose cough intensity decreased to 2/10 in VAS; solid line: patients whose cough intensity decreased to 5/10 in VAS VAS, visual analogue scale

**Figure 4.**
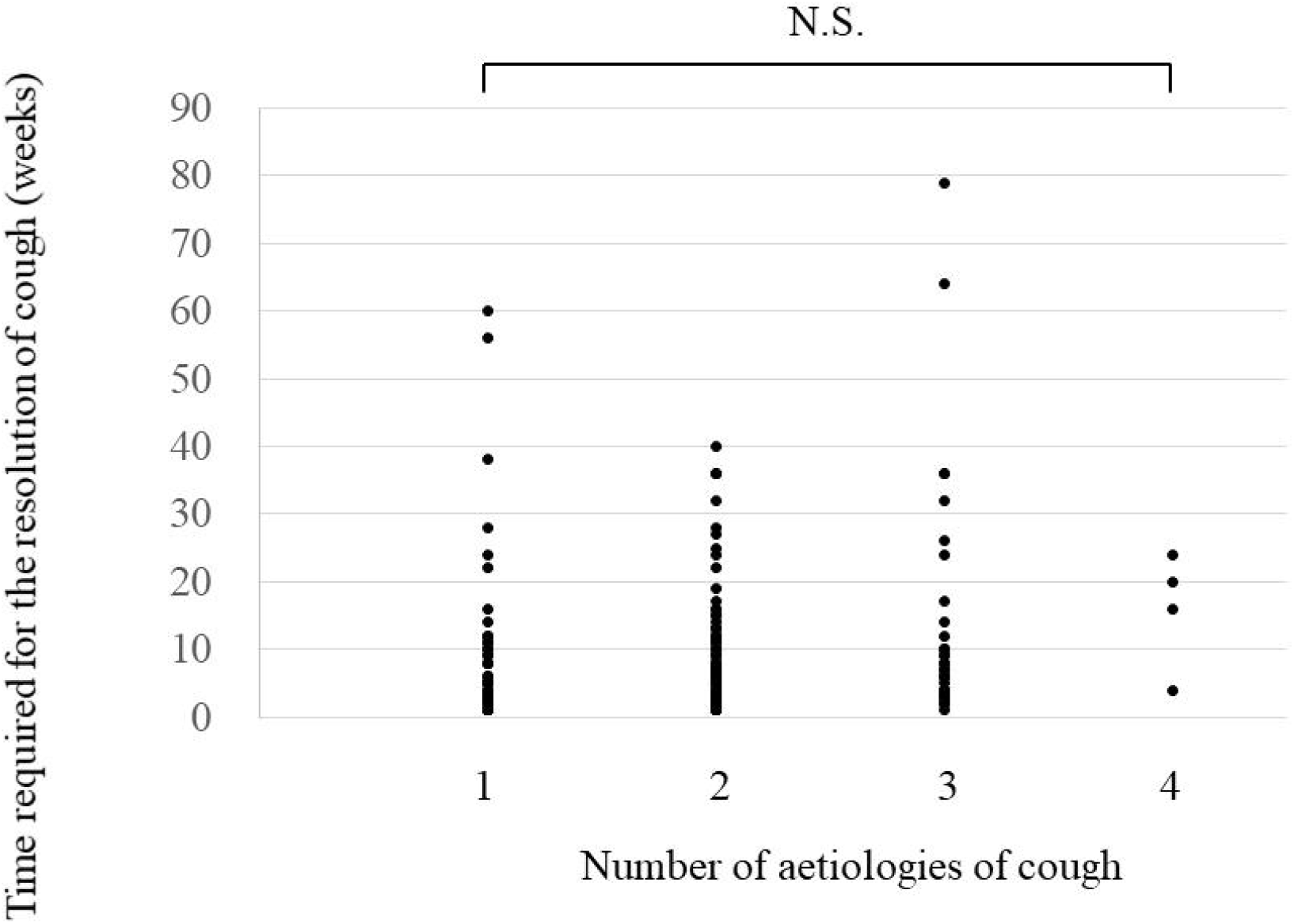
Comparison of cough resolution periods among patient groups with different number of cough aetiologies N.S: not significant

**Table 4.** Aetiologies of chronic cough diagnosed using both the pathophysiological diagnostic procedure and treatment response procedure in 291 patients

|  |  |
| --- | --- |
| Single cause | 72 |
| AC | 7 |
| CVA | 60 |
| CPA | 4 |
| MISB | 1 |
| Dual causes | 160 |
| CVA + SBS | 81 |
| AC + CVA | 49 |
| CPA + SBS | 10 |
| AC + SBS | 8 |
| CVA + GER | 4 |
| CVA + MISB | 3 |
| AC + CPA | 1 |
| CPA + MISB | 1 |
| CPA + DPB | 1 |
| CPA + GER | 1 |
| SBS + Bronchorrhea | 1 |
| Tiple causes | 53 |
| AC + CVA + SBS | 37 |
| AC + CVA + GER | 6 |
| CVA + SBS + MISB | 4 |
| SBS + CPA + MISB | 3 |
| AC + SBS + CPA | 1 |
| AC + CVA + MISB | 1 |
| AC + CVA + DPB | 1 |
| Quad causes | 6 |
| AC + CVA + GER + SBS | 4 |
| AC + CVA + SBS + MISB | 2 |
| Unknown | 0 |

AC: atopic cough; CPA: cough predominant asthma; CVA: cough variant asthma; DPB: diffuse panbronchiolitis; GER: gastroesophageal reflux; MISB: mucoid impaction of small bronchi; SBS: sinobronchial syndrome

## Discussion

The present study showed that our pathophysiological diagnostic procedure resulted in a high success rate of diagnosis and treatment of CC. No studies using therapeutic diagnostic procedures have included the results of treatment for CC; thus, this is the first report concerning the results of treatment for CC.

In the present study, 97.3% of patients who continued treatment achieved complete cough elimination. We believe that this outcome of treatment is outstanding considering that existing studies have evaluated cough treatment efficacy as improvement in various measures, not as cough elimination; and because *unexplained* CC is diagnosed in up to 46% of patients referred to specialty cough clinics ^20^. Generally, cough guidelines have shown that the period of initial treatment for a suspected cough aetiology was 1–8 weeks ^21-23^. In a self-perpetuating cycle, cough itself damages the airways, which results in the exacerbation of airway inflammation and cough ^24-26^. Therefore, accurate pathophysiological diagnosis and sequential rapid treatment may have improved the therapeutic effect in this study.

In the present study, rwo or more aetiologies were diagnosed in 72.7% of patients with CC. In these cases, use of therapeutic diagnostic procedures is more difficult and time-consuming. Guidelines indicate the limited accuracy of therapeutic diagnostic procedures in CC is with multiple aetiologies ^23^. We believe that cases with multiple aetiologies accounted for such a large proportion of all cases because we used the pathophysiological diagnostic procedure and investigated the true diagnosis when cough completely resolved. Elderly patients had a higher number of aetiologies than did non-elderly patients (p =0.020). Patients in our study were older than those in previous studies ^27-29^, and aging may influence findings to some extent. Among our 300 patients,

AC, CVA, and SBS were the major causes of CC in 256, 149, and 119 patients, respectively. In 2005, we reported that AC, CVA, and SBS were the three major causes of CC in the Hokuriku region of Japan ^27^. Therefore, the present study showed that, at least in our institution, the causes of CC remained unchanged over the past 15 years, during which period the publications of two guidelines for cough management ^3, 30^ and advances in cough research and diagnostic procedures occurred.

To the best of our knowledge, there have been few reports concerning the treatment period required for complete resolution of cough. One study from China showed that cough improvement (complete resolution or partial improvement) in a tertiary clinic required >14 weeks ^31^. In the study, cough completely resolved in 65 of 155 (41.9%) patients in tertiary care clinics and 67 of 193 (34.7%) patients in secondary care clinics, and cough improved by at least 50% in 76 of 155 (49.0%) patients in tertiary care clinics and 97 of 193 (50.3%) patients in secondary care clinics. Two retrospective cohort studies examined longitudinal CC prognosis. Kang et al. showed that 64 of 323 patients with CC (19.8%) had persistent cough 4 years after assessment and treatment in a Korean tertiary clinic ^32^. Koskela et al. showed that 31 of 68 patients with CC (46%) had continued regular cough 5 years after the initial assessment at Finnish University. These two studies, however, did not show detailed causes of cough. In our study, the median time required for complete resolution of cough was 5.0 weeks (95% CI: 4.3∼5.7), and the number of aetiologies did not influence the time required for complete cough resolution. Because patient characteristics, assessment methods, and treatment strategies used in the present study are different from those of previous studies, prognoses are not comparable. However, this study suggests that assessment and treatment of CC based on pathophysiological diagnosis may improve the prognosis compared to that of CC existing methods.

ICSs are principal drugs used for the treatment of eosinophilic airway diseases, such as CVA, AC, and CPA. CVA may cause bronchial asthma and future airway remodelling, in requiring long-term ICS treatment. On the contrary, AC does not cause bronchial asthma or airway remodelling;,therefore, ICS treatment can be discontinued when the cough is resolved. We must pathophysiologically differentiate CVA from AC, with their different prognoses, before starting treatment; we fear that the empirical use of ICS recommended by some guidelines ^21-23^ will make it impossible to distinguish between CVA and AC. Long-term ICS use is associated with an increased risk of pneumonia ^33^ and mycobacterial infection ^34, 35^ in patients with chronic obstructive pulmonary disease. There is no direct evidence of risk of using ICS in patients with CC, but the routine use of ICSs in patients with cough should be avoided.

In summary, our study demonstrated the usefulness of pathophysiologic diagnostic procedures for determining aetiologies of CC. A recent study reported that ineffectiveness of treatment and unclear diagnosis were major unmet needs in CC ^36^. We recommend advancing from therapeutic to pathophysiologic diagnostic procedures.

## Conclusion

Therapeutic diagnostic procedures for CC require at least a one-week treatment period, and a further treatment period would be necessary in case of treatment failure. Furthermore, when two or more causes are present,, the diagnosis may be further complicated. In contrast, pathophysiologic diagnostic procedures require merely two days to measure cough receptor sensitivity to inhaled capsaicin and cough response to methacholine-induced bronchoconstriction before initiation of treatment. Thus, a pathophysiology-based diagnostic approach should be employed globally, which may improve the management of patients with CC.

## Data Availability

The data that support the findings of this study are available from the corresponding author, Johsuke Hara, upon reasonable request.

## Acknowledgements

We would like to thank Editage (https://www.editage.jp/) for the English language review.

This research did not receive any specific grant from funding agencies in the public, commercial, or Units.

